# From a Voxel to Maps: A Comparative Study of sLASER SVS and 3D-CRT-FID-MRSI at 3 T and 7 T

**DOI:** 10.1101/2025.08.27.25334335

**Authors:** Zeinab Eftekhari, Korbinian Eckstein, Bernhard Strasser, Lukas Hingerl, Fabian Niess, Wolfgang Bogner, Thomas B Shaw, Markus Barth

## Abstract

Magnetic resonance spectroscopy (MRS) enables non-invasive assessment of brain metabolites and is commonly implemented using single-voxel spectroscopy (SVS) or magnetic resonance spectroscopic imaging (MRSI). This study directly compares the reproducibility of SVS-based semi-localization by adiabatic selective refocusing (sLASER) and 3D-Concentric Ring Trajectory-based Free Induction Decay MRSI (3D-CRT-FID-MRSI) at 3T and 7T in the same cohort of healthy participants. To explore MRSI’s capabilities for regional metabolite quantification and reproducibility assessment, three masking strategies were applied. Additionally, two spatial averaging approaches for MRSI, averaging before vs. after spectral fitting, were evaluated. Coefficients of variation (CV) and voxel-wise correlation analyses were used to assess intra-subject and inter-session reproducibility. Results showed good-to-excellent reproducibility across both techniques, with SVS generally providing lower CVs at 7T, while MRSI outperformed SVS in several metabolites at 3T. MRSI allowed tissue-specific analysis, with lower CVs observed in WM compared to GM, especially at 7T. Although MRSI reproducibility was slightly reduced at 7T likely due to longer scan times and lack of prospective motion correction, the spatial coverage and retrospective region analysis makes it an attractive alternative to SVS for many brain regions. This study demonstrates that both sLASER and CRT-FID-MRSI provide reproducible metabolite measurements at 3T and 7T. The findings highlight MRSI’s advantages for retrospective multi-regional and tissue-specific analysis, facilitating its integration into future clinical research.

## 1. Introduction

Magnetic resonance spectroscopy (MRS) is a non-invasive tool that provides a window into brain metabolism, helping researchers and clinicians understand both normal biochemical processes and the changes that occur in diseases. Two main approaches dominate the field: single-voxel spectroscopy (SVS)^1^ and magnetic resonance spectroscopic imaging (MRSI)^2^. While both techniques aim to quantify brain metabolites, they have unique strengths and applications. SVS captures information from a single region of interest (ROI) within the brain, making it highly precise and reliable for targeted metabolite studies^3-5^. However, it cannot provide information of spatial heterogeneity in metabolite concentrations, which is relevant for studying many whole-brain diseases. SVS also captures a mix of multiple tissue types, leading to partial volume effects^6-9^. A challenge in MRS in general is its sensitivity to static magnetic field strength (B_0_) inhomogeneities within the volume of interest (VOI), requiring careful corrections via time-consuming shimming procedures to improve spectral resolution and quantification accuracy^10^.

In contrast, MRSI can create maps of metabolite concentrations across larger regions of the brain. This allows researchers to study metabolic variabilities, such as differences in glutamate (Glu) or glutamine (Gln) levels between different brain areas, as well as their alterations in pathological conditions such as brain tumours and multiple sclerosis (MS)^11,12^. The smaller voxel sizes in MRSI have the advantage of reduced intra-voxel B_0_ inhomogeneities at the single-voxel level, but this is partially compensated by the fact that MRSI requires shimming over a much larger VOI and hence intrinsically more inhomogeneous volume than SVS^13-15^. Other MRSI-specific challenges include lower SNR per voxel, longer scan times, and susceptibility to imaging artifacts^16,17^. Recent innovations, such as free induction decay based-MRSI using a concentric ring trajectory (CRT-FID-MRSI) acquisition, have successfully enhanced spatial resolution and data quality^14,18-21^. CRT-FID-MRSI uses fast and SNR-efficient non-Cartesian spatial-spectral encoding with high spatial resolution, which makes high-quality metabolite mapping within clinically practical scan times possible^17^. FID-MRSI captures the FID signal directly without a spin-echo, thereby eliminating the need for additional refocusing pulses, which enables an ultra-short acquisition delay (∼1.3 ms)^14,22-25^. This feature minimizes TLJ decay and eliminates J-evolution-related SNR loss, which is a major challenge at ultra-high fields (UHF). This improves quantification accuracy for short-TLJ, and especially J-coupled metabolites^14,22-25^. By avoiding refocusing pulses, FID-MRSI reduces the specific absorption rate (SAR), which is particularly advantageous at high B_0_, where SAR constraints are more stringent^6,14^. As a result, this technique remains within 60% of the SAR limit even at 10.5T in humans^26^. Additionally, FID-based MRSI is inherently less sensitive to transmit radiofrequency field (B_1_^+^) inhomogeneities, further improving spectral quality at ultra-high fields^14,19^.

At higher B_0_, such as 7T, both SVS and MRSI benefit from enhanced spectral resolution and SNR^27^. These improvements make it easier to separate overlapping peaks, such as those of Glu and Gln, which are difficult to distinguish at lower B_0_ ^28^. However, 7T also introduces challenges, including greater chemical shift displacement errors (CSDE) and increased sensitivity to inhomogeneous B_1_ ^+^ fields^27^. Advanced sequences like semi-localization by adiabatic selective refocusing (sLASER)^29^ for SVS and CRT-FID-MRSI^14^ for spectroscopic imaging have been developed to address these issues.

This study compares the reproducibility of sLASER^30^ and CRT-FID-MRSI^14^ at 3T and 7T. By scanning the same group of healthy participants twice about one week apart acquiring both SVS and MRSI. We acquired SVS in two brain regions (precentral gyrus and paracentral lobule), which are highly relevant to motor function and key areas affected in related pathologies such as motor neuron disease (MND), the focus of our future research.

Moreover, this study aims to evaluate MRSI performance at 3T and 7T, explore advanced MRSI data analyses like region-specific masking, and assess how different region-averaging methods, specifically fitting before averaging (“fit-first”) versus averaging before fitting (“average-first”), impact reproducibility and quantification accuracy particularly for low-concentration metabolites.

## 2. Materials and Methods

### 2.1. Hardware

This study was conducted at 3T (PrismaFit MR scanner; Siemens Healthineers, Erlangen, Germany) using a 64-channel receive-only head coil in combination with the integrated body transmit coil and at 7T (MAGNETOM 7T Plus MR scanner; Siemens Healthineers, Erlangen, Germany), with a 1Tx/32Rx head coil (Nova Medical, Wilmington, MA, USA).

### 2.2. Human Subjects

Five healthy volunteers (aged 25–33 years; mean age = 29 years, SD = 2.8; 3F/2M) underwent two scanning sessions at both 3T and 7T, with an interval of five to nine days between measurements. All participants provided written informed consent prior to the study, which was conducted in accordance with the ethical standards set by the local Human Research Ethics Committee.

### 2.3. Data Acquisition and Sequence Parameters

All participants underwent an anatomical T_1_-weighted MP2RAGE scan for voxel positioning and tissue segmentation^31^. The scanning parameters were as follows: at 7T, voxel size = 0.8×0.8×0.8 mm³, TR = 4300 ms, TE = 2.4 ms, TA = 6:54 mins, FA = 5°, matrix size = 192×224×256; at 3T, voxel size = 0.9×0.9×0.9 mm³, TR = 1900 ms, TE = 2.3 ms, TA = 4:10 mins, FA = 9°, matrix size = 176×240×256^31^.

#### 2.3.1. SVS

Before acquiring SVS data, B_0_ shimming was performed within the VOI using 1^st^ and 2^nd^ order shimming via FAST(EST)MAP^32,33^ across all subjects and sessions consistently with one linear iteration, one all shim iteration and one additional linear iteration. RF transmitter power and water suppression using variable pulse power and optimised relation delays (VAPOR) were calibrated and was interleaved with outer volume suppression (OVS) pulses to suppress unwanted coherences, ensuring consistent flip angles and water suppression across all VOIs^34^. All sLASER spectra were acquired using the following parameters: At 7T: voxel size = 25×25×25 mm³, TR = 8000 ms, TE = 26 ms, 16 averages, TA = 2.5 mins At 3T: voxel size = 25×25×25 mm³, TR = 2000 ms, TE = 28 ms, 64 averages, TA = 2.5 mins^29^. A longer TR was used at 7T due to stricter SAR limits. sLASER reduces CSDEs by using high-bandwidth gradient offset independent adiabatic (GOIA)-WURST adiabatic full passage (AFP) refocusing pulses (45kHz), resulting in CSDEs of 0.27 % per ppm at 3T and 0.66% per ppm at 7T ensure accurate voxel localization^5,29,35^ and the slice-selective excitation pulses with bandwidth of 4kHz at 3T and 6kHz at 7T, leading to CSDEs 3% and 4.9% per ppm, respectively. Non-suppressed water spectra with matching parameters were acquired for eddy current correction and water referencing (VAPOR and OVS schemes turned off). Two brain regions within the motor cortex of the right hemisphere—the precentral gyrus (upper limb motor homunculus region) and the paracentral lobule (lower limb motor homunculus region)—were chosen as these are areas targeted in our future clinical research (Figure 1). To ensure consistency, technologists adhered to strict voxel placement protocols, including overlaying the original anatomical scan with the follow-up scan, and positioning based on printouts of the initial visit’s voxel positioning during follow-up scans.

**Figure 1:**
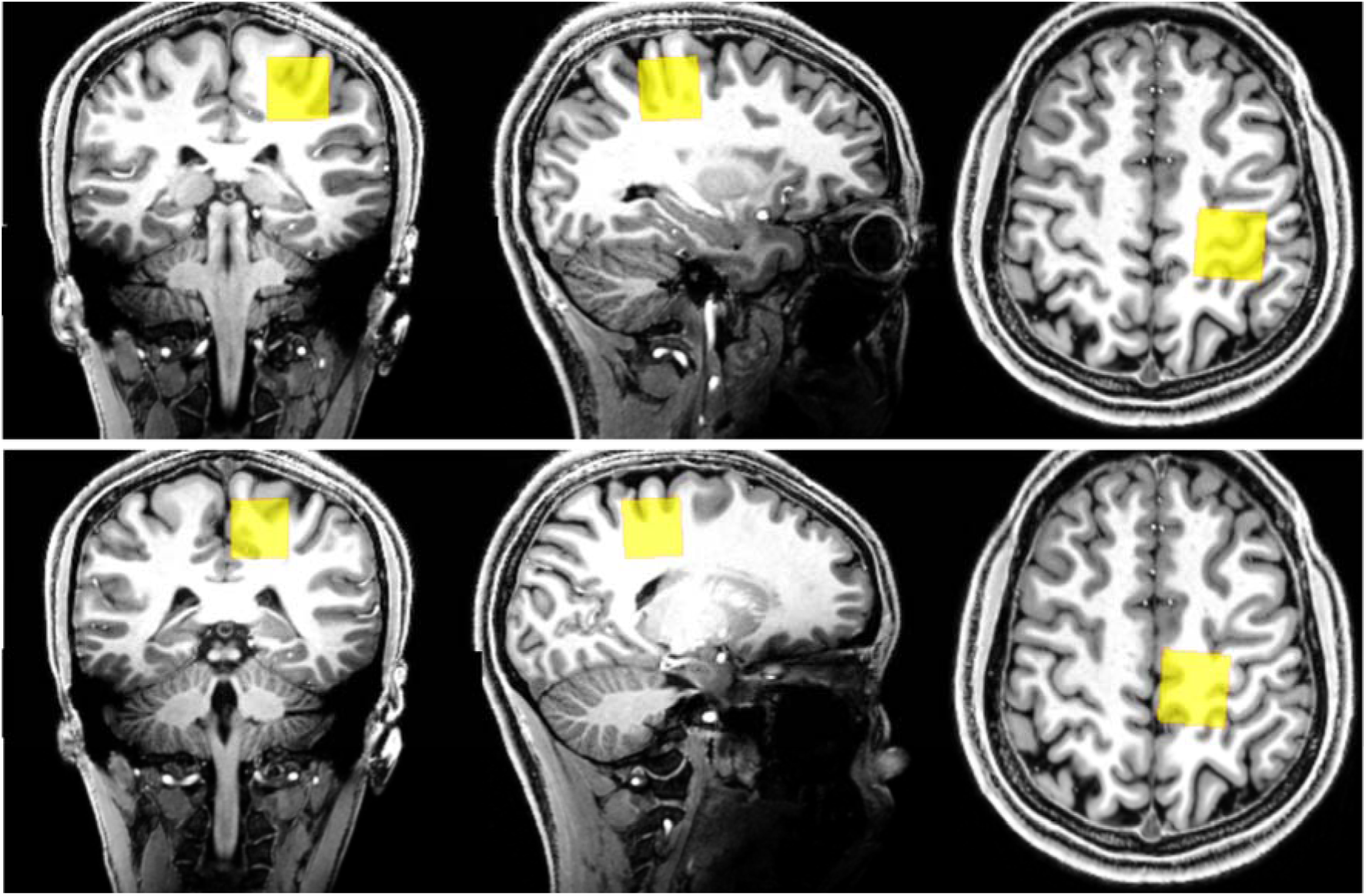
Volumes of interest (VOIs) positioned for single voxel spectroscopy in the precentral gyrus (top; upper limb) and the paracentral lobule (bottom; lower limb) shown on T_1_-weighted images acquired at 7T.

#### 2.3.2. CRT-FID-MRSI

Details regarding the sequence design and implementation are thoroughly outlined in Hingerl et al^19^. At 7T, the protocol included B_1_^+^ mapping for flip-angle optimization^36^. At both 3T and 7T, B_0_-shimming for 1^st^ and 2^nd^ order spherical harmonics B_0_ shimming was performed terms over the MRSI slab. At 3T, an additional interleaved volumetric navigator was available for real-time motion and shim correction^37^. 3D-CRT-FID-MRSI parameters were: 7T: voxel size = 3.4×3.4×3.5 mm³, TR = 460 ms, TE = 1.3 ms, TA = 12:11 mins, FA = 39° (calculated as the nominal average Ernst angle for NAA, tCr, tCho, Glu, and mIns), matrix size = 64×64×31, FOV = 220×220×110 mm³, Readout duration = 311 ms, spectral bandwidth = 2778 Hz; 3T: voxel size = 6.3×6.3×6.2 mm³, TR = 950 ms, TE = 0.8 ms, TA = 4:14 mins, FA = 70°, matrix size = 32×32×21, FOV = 200×200×130 mm³, Readout duration = 441 ms, spectral bandwidth = 1325 Hz^20,36,38^. Utilizing a short, high bandwidth slice-selective excitation pulse, reducing CSDE in the slice direction to approximately 5% per ppm at 7T^14^. At 7T variable temporal interleaves (1–3, depending on ring radii) to maintain spectral bandwidth for larger readout circles and reduce scan times, and optimised water suppression enhanced through T_1_ effects (WET) were used^19,36^. A slab thickness of 60 mm, parallel to the horns of the corpus callosum, was selected for MRSI acquisitions. We aimed for roughly similar SNR at 3T and 7T and thus increased resolution at 7T. These parameters were designed to optimize spatial resolution, acquisition efficiency, and spectral quality^20,37^.

### 2.4. Data Processing

All raw data were stored in Siemens format for offline post-processing.

#### 2.4.1. SVS

All SVS post-processing steps were automated using Osprey (v. 2.5.0), an open-source MRS analysis toolbox, based on the MATLAB platform (MATLAB R2022a, MathWorks, Natick, MA, USA) with the integrated LCModel (linear-combination model) fitting algorithm and Statistical Parametric Mapping’s toolbox (SPM12)^39-41^. The automated pipeline in Osprey is composed of several critical steps to ensure data quality and accurate spectral analysis^40^. These steps included:

a. combining raw data from multiple coils for optimal SNR,
b. correcting frequency and phase drifts in individual shots,
c. compensating for eddy-current effects using the water reference scan,
d. averaging the spectra,
e. removing residual water signals, and
f. applying baseline correction.

Additionally, grey matter (GM), white matter (WM), and cerebrospinal fluid (CSF) tissue segmentation was conducted using SPM12. Key quality metrics, such as the SNR of total creatine (tCr) and the full width at half maximum (FWHM) of the water peak (ppm), were calculated from the post-processed spectra to assess data quality.

#### 2.4.2. CRT-FID-MRSI

MRSI data were processed offline using a custom-designed software pipeline^42^, integrating MATLAB (MATLAB R2022a, MathWorks, Natick, MA, USA), Bash (v5.2.21, Free Software Foundation, Boston, MA, US), FSL (Analysis Group, FMRIB, Oxford, UK^43^), and MINC tools (McConnell Brain Imaging Centre, Montreal, QC, Canada^44^). The pipeline included the following steps:

a. coil combination using iMUSICAL (interleaved multichannel spectroscopic data combined by matching image calibration data), which is based on interleaved water calibration scans for optimal spectral consistency^21,45^;
b. k-space reconstruction employing in-plane convolution gridding, which weights non-Cartesian sampling points to align with the Cartesian grid^21,46^;
c. correction for the time delay of acquisition samples using off-resonance compensation^47^;
d. spatial filtering with a Hamming filter to improve the quality of metabolite maps.
e. Additionally, lipid signals were reduced post-measurement using L2 regularization to minimize contamination in metabolite quantification^48^.

While corrections for B_1_ inhomogeneities were not applied, prior studies have demonstrated that local flip angle variations exert minimal effects on T_1_ weighting for FID-MRSI^49^. These processing steps ensure robust spectral reconstruction and minimized artifacts while maintaining the integrity of the metabolic information (Figure 3)^42^. SNR for NAA was estimated voxel-wise using the pseudo-replica method, which incorporates receiver noise prescans^22^. Additionally, the FWHM of the spectral peaks for tCr at 3.02 ppm was calculated from the LCModel fits to evaluate spectral resolution and line-shape quality.

**Figure 2:**
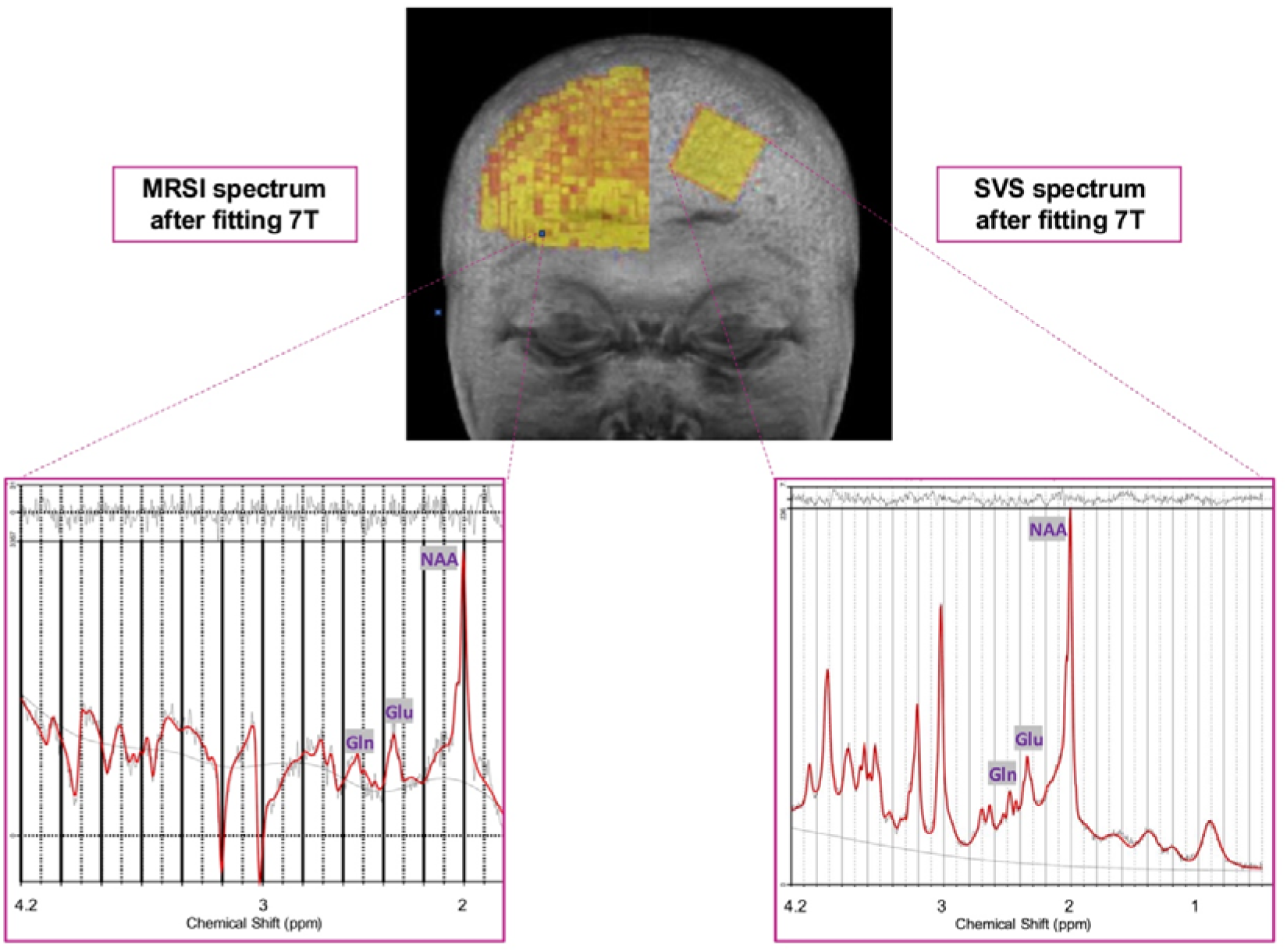
Representative LCModel fits of ^1^H MR spectra, and voxel sizes from one subject measured at 7T using both techniques are displayed. The left panel shows an MRSI spectrum, while the right panel displays an SVS spectrum. This figure highlights the significant difference in spatial resolution between the two techniques: the SVS voxel size is 25×25×25 mm³, encompassing a large brain region, whereas the MRSI voxel size is much smaller at 3.4×3.4×3.5 mm³, allowing for higher spatial detail. It also highlights the differences in the spectra of these two methods, namely multiplet peaks of glutamine (Gln) and the small peak of *N*-acetylaspartylglutamate (NAAG). N.B.: the distinct phasing of the MRSI spectra (e.g., choline (Cho) and creatine (Cr) flipped peaks) is due to the 1.3 ms acquisition delay and the absence of first-order phase correction, which is inherent to the acquisition scheme, but is appropriately considered during the LCModel fitting.

**Figure 3:**
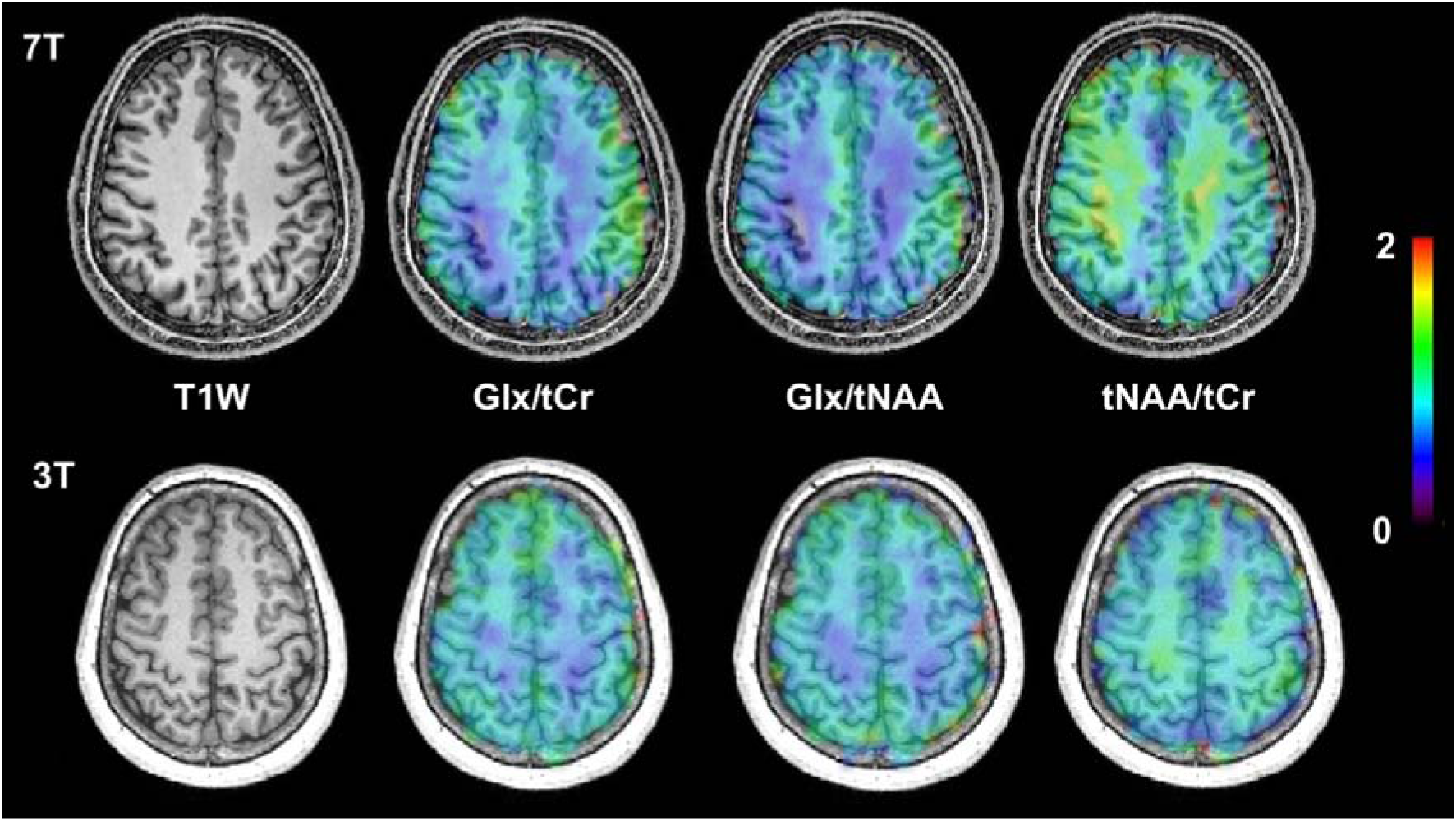
Representative metabolic maps of three important metabolites [tNAA =*N*-acetylaspartate (NAA)+ *N*-acetylaspartyl (NAAG); Glx= glutamate (Glu) + glutamine (Gln); tCr= creatine (Cr) + phosphocreatine (PCr) obtained with 3D-MRSI overlaid on anatomical T_1_-weighted images acquired at 7T (top) and 3T (bottom). These illustrate the ability of 3D-CRT-FID-MRSI to generate high-resolution maps. Colour from cold to warm indicate higher relative concentrations. The maps are interpolated for visualization. All metabolic maps are reported in arbitrary units.

### 2.5. Spectral Fitting and Quantification

The analysis of ^1^H-MR spectra were conducted using LCModel^39^. For SVS, the chemical shift range of 0.5 to 4.2 ppm was analysed^50^, while for CRT-FID-MRSI, the range was limited to 1.8 to 4.2 ppm to exclude the lipid resonance at 1.3 ppm (Figure 2). To mitigate lipid contamination in 3T SVS, a further exclusion gap of 1.1 to 1.85 ppm was applied. The LCModel basis sets for 3T and 7T included 19 simulated metabolites and a measured macromolecule spectrum as previously reported^18,37,51-53^. tCr served as the internal concentration reference for both SVS and MRSI.

### 2.6. Masking Strategies for Data Analysis

To enable meaningful comparisons and to fully utilise the unique capabilities of MRSI, separate masking strategies were considered and evaluated in this study (Figure 4). These approaches allowed for the extraction of metabolite data from specific ROIs tailored to different analytical objectives. These distinct masking approaches were: SVS-based masking (lower limb and upper limb region), tissue-specific masking (GM and WM), and atlas-based masking (precentral gyrus and paracentral lobule). Each mask, whether derived from the SVS voxel, tissue segmentation, or anatomical atlas, was resampled to match the resolution of MRSI metabolite maps using advanced normalisation tools (ANTs) transformations with an identity transform, binarising them and then overlaid to MRSI neurochemical maps^43,54,55^. This process ensured that voxel-wise comparisons were performed in spatially congruent regions between techniques and repeated measurements. Additionally, T_1_-weighted images for each subject from session one was registered to a localiser image from session two.

**Figure 4:**
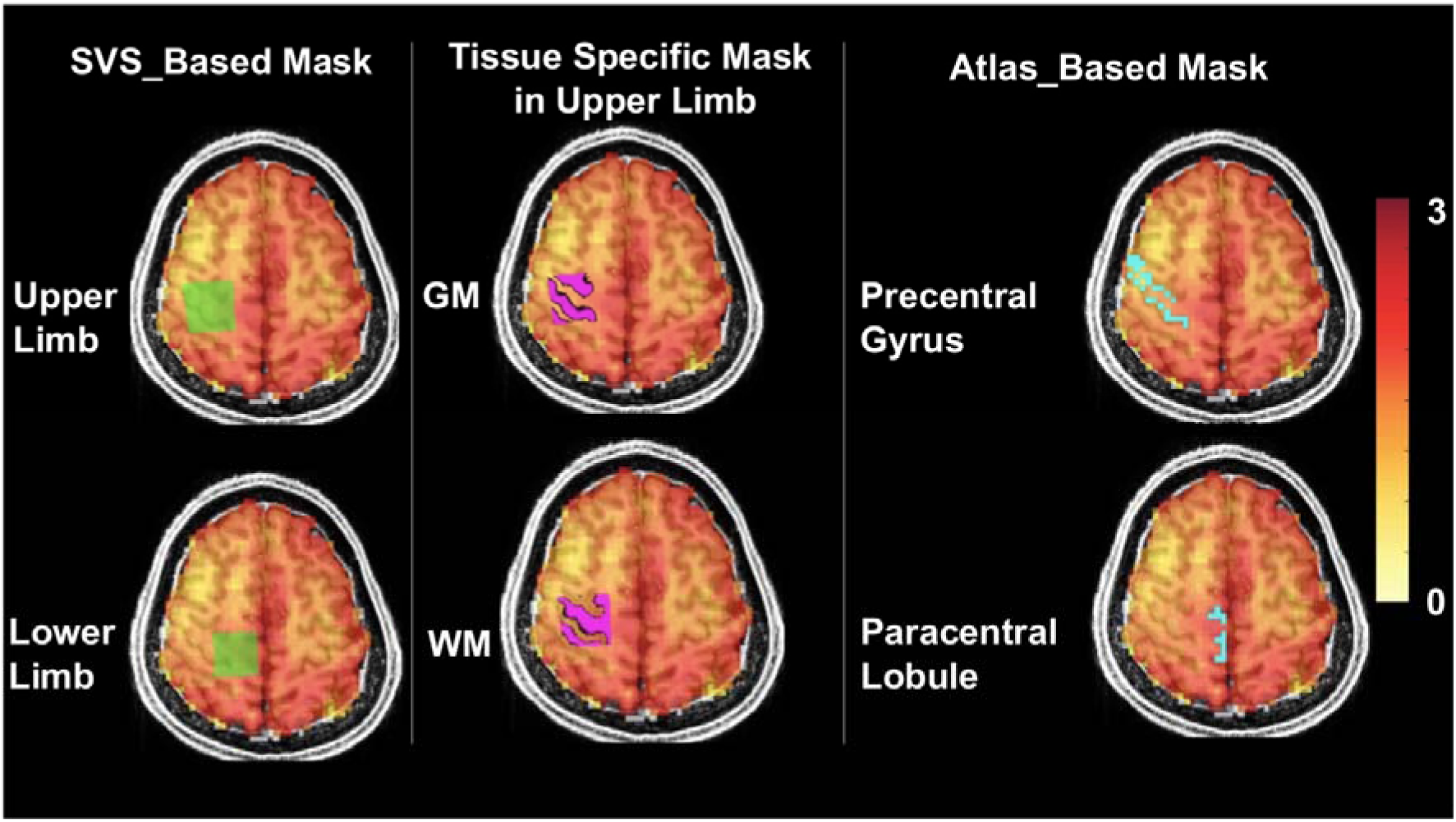
An example of masking strategies applied to a single representative subject at 7T using their MP2RAGE anatomical image as the base layer, with the hot overlay representing the total *N*-acetylaspartate/total creatine (tNAA/tCr) MRSI map. Left: voxel masks derived from sLASER sequences in the upper and lower limb regions of the motor cortex (green). Middle: grey matter (GM) and white matter (WM) segmentations within the upper limb SVS mask, generated via SPM12 (pink). Right: Atlas-based masks using Fastsurfer highlighting the paracentral lobule and precentral gyrus (blue). All masks extend in 3D.

#### 2.6.1. SVS-Based Masking

Masks were overlaid on the SVS voxel position for each session and were applied to the MRSI neurochemical maps. This ensured that ROIs for both techniques were spatially aligned.

#### 2.6.2. Tissue-Specific Masking: GM and WM

To leverage MRSI’s ability to capture metabolite concentrations within distinct tissue types, additional masks were generated to isolate GM and WM within the SVS voxel using SPM12. Only masks for GM and WM with consistent voxel overlap across sessions were identified using minimum overlap masks.

#### 2.6.3 Atlas-Based Masking

All T_1_-weighted images were processed using FastSurfer for brain structure segmentation^56^. Masks for the precentral gyrus and paracentral lobule corresponding to the motor cortex were extracted, due to their relevance to our future clinical research.

### 2.7. Region Averaging Approaches for MRSI Data Processing

For MRSI data, two region-based quantification strategies were employed: 1) “fit-first” and 2) “average-first”. This comparison aimed to evaluate whether noise reduction from spectral averaging prior fitting could improve the quantification of low-concentration metabolites such as GABA, Gln, and Tau, which are particularly sensitive to SNR and fitting robustness.

#### 2.7.1. “Fit-First” Approach

In the “fit-first” method, metabolite concentration maps were obtained by voxel-wise spectral fitting using LCmodel. After fitting, the metabolite values were averaged over region-specific masks (e.g., SVS-based, GM/WM, or atlas-based masks). This simple approach enabled straightforward computation of average metabolite concentrations within the masked region. It preserves the detailed spatial information provided by MRSI. Researchers can analyse individual voxels, identify outliers, and exclude poor-quality data, ensuring better quality control.

#### 2.7.2. “Average-First” Approach

The “average-first” method focuses on improving spectral quality before fitting. The steps include:

1. **Mask Application**: Select only the spectra within the masked region.
2. **Frequency and Phase Correction**: B_0_ and B_1_ inhomogeneities introduce local variations in the magnetic field^13^. To align all spectra before averaging, each spectrum were aligned for phase by internal reference (division by third time point) and for frequency by applying frequency shifts based frequency shifts determined by LCModel.
3. **Averaging**: All corrected complex spectra within the ROI were averaged to generate a single, high-SNR spectrum representing the region.
4. **Spectral Fitting**: The averaged spectrum was fitted using LCModel to quantify metabolite concentrations.

### 2.8. Statistical Analysis

A total of 40 SVS scans (2 voxels × 2 sessions × 5 subjects × 2 fields) and 20 MRSI scans (2 sessions × 5 subjects × 2 fields) were collected. Metabolite estimates with Cramér-Rao lower bounds (CRLBs) exceeding 50% were excluded to ensure accurate quantification in line with previous literature to avoid overly aggressive rejection of low-concentration metabolites^57^. For metabolites prone to strong negative correlations (r < −0.5), such as PCho and GPC, and NAA and NAAG combined sums were reported to simplify analysis.

The reproducibility of metabolite quantification between SVS and MRSI was assessed using CV analysis. CVs were calculated for metabolite ratios to tCr, demonstrated by Wijtenburg et al., 2018, to evaluate reproducibility at both 3T and 7T field strengths^18,58-60^. Lower CV values indicate higher reproducibility, reflecting minimal variability in repeated measurements.

To evaluate the agreement of MRSI between sessions one and two for each metabolite at each field strength, voxel-wise analyses were performed using metabolite ratio maps from both scanning sessions covering the entire VOI while excluding non-brain voxels. This approach goes beyond regional averaging by examining the spatial consistency of metabolite distributions across the whole brain, providing a more comprehensive assessment of reproducibility. Following the application of minimum overlap masks, the 3D maps were flattened into a 1D representation to facilitate direct voxel by voxel comparisons. Correlation coefficients (Pearson’s r) were computed for metabolite concentrations across the two sessions to assess how well the metabolite concentrations matched between sessions and highlighting how consistently the metabolic patterns were reproduced.

### 2.9. Data and Code Availability

The full set of scripts and analysis pipelines utilized in this study, as well as the processed CSV files containing metabolite concentration data from MRS and MRSI, are publicly accessible via the GitHub repository [https://github.com/Zeinabeftekhari/MRSI_SVS_comparison.git].

## 3. Results

### 3.1. Comparison Between MRSI and SVS techniques

#### 3.1.1. Data Quality of Measurements

Figure 2 illustrates representative spectra obtained using both MRSI and SVS, along with their LCModel fits and residuals. No averaged data were excluded from the analysis except GABA at 7T using MRSI which showed CRLB higher 50% for the whole region inside the masks for 16/ 20 datasets. For MRSI, the mean SNR for NAA and FWHM of tCr (ppm) were calculated using the “fit-first” region averaging method within the SVS-defined masks at each field strength. Table 1 presents the sLASER tCr SNR and water FWHM (ppm) at both fields, while Figure 5 provides the corresponding values for MRSI. For MRSI SNR, no significant differences between 3T and 7T were found for either the lower limb (3T: 18.75 ± 2.43; 7T: 19.80 ± 3.04; *p =* 0.408) or the upper limb (3T: 19.91 ± 3.81; 7T: 21.97 ± 3.53; *p =* 0.229) regions, suggesting that signal quality per voxel remains consistent across field strengths in these regions using our proposed protocol setup. MRSI FWHM was similar at 3T and 7T (0.02 ppm or 0.03 ppm), but a significant difference between 3T and 7T was observed for the lower limb region (3T: 0.028 ± 0.006; 7T: 0.038 ± 0.007; *p* = 0.004), with higher FWHM values at 7T. However, no significant difference was found for the upper limb region (3T; 0.033 ± 0.007, 7T; 0.036 ± 0.005, *p* = 0.350).

**Figure 5:**
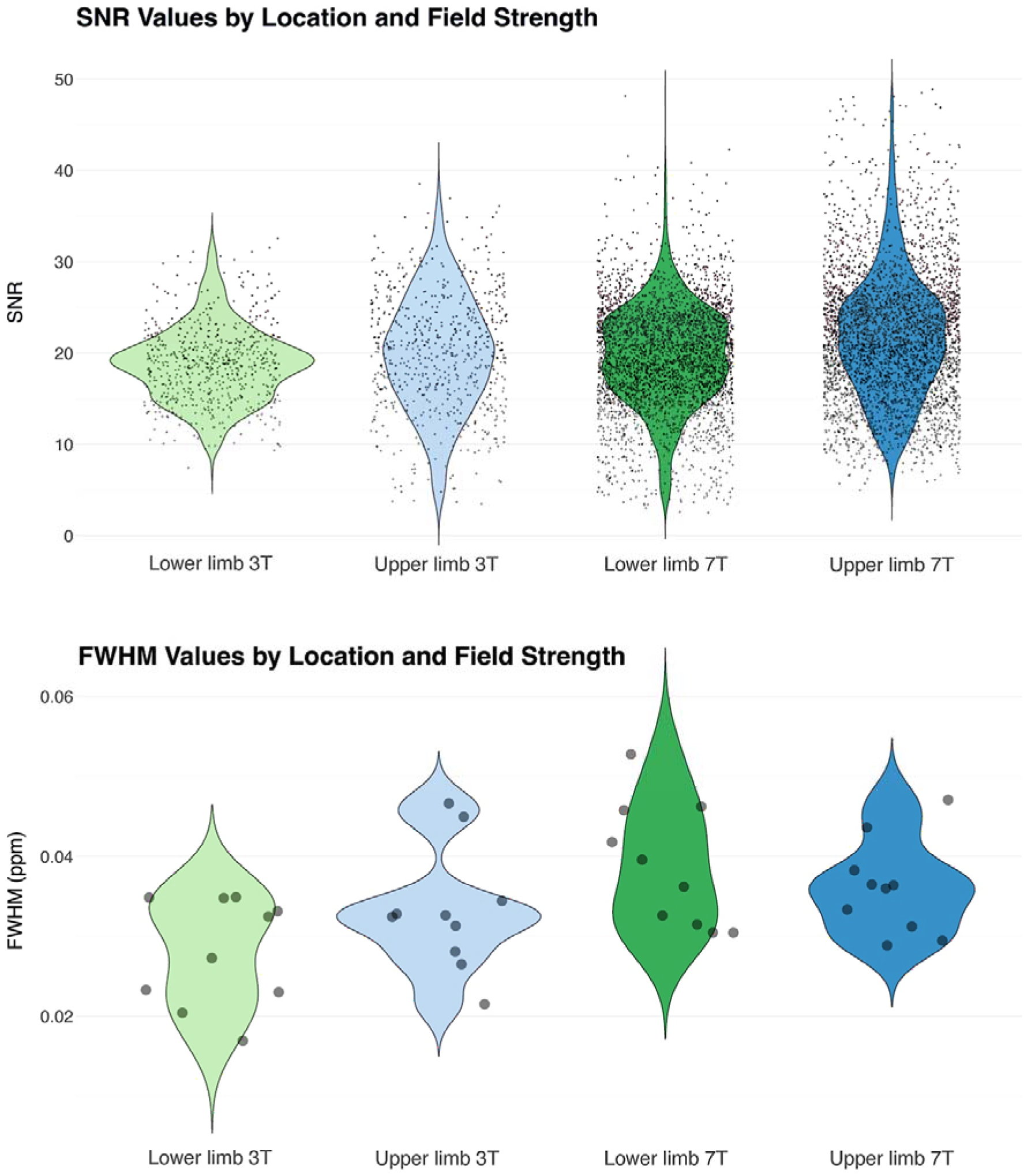
Violin plots illustrate *N*-acetylaspartate (NAA) signal-to-noise (SNR) values (top) for MRSI as black dots for individual voxels within the SVS masks, collected from five subjects over two sessions. The lower limb region is represented in light green and dark green, while the upper limb region is depicted in light blue and dark blue. 7T has more voxels due to the higher resolution/smaller voxel size (64×64×31 compared to 32x32x21). We aimed for roughly similar SNR at 3T and 7T and thus increased resolution at 7T. Displaying the distribution of SNR values per voxel rather than an average better reflects the heterogeneity in signal strength across the SNR masks. The bottom violin plot depicts the MRSI full width at half maximum (FWHM) values of total creatine (tCr) averaged over all voxels obtained from two scans of five subjects. The lower limb region is represented at 3T in light green and at 7T in dark green, while the upper limb region is shown at 3T in light blue and at 7T in dark blue.

**Table 1:**
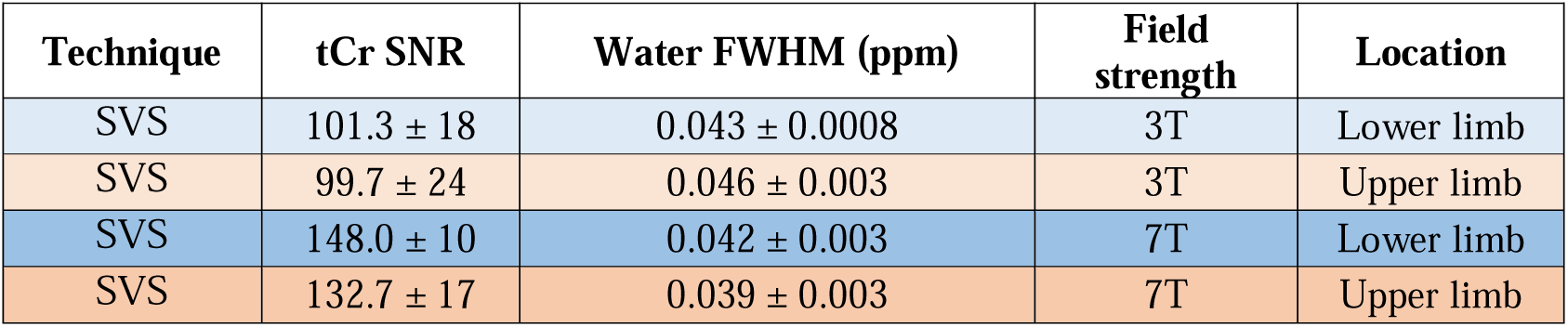
Total creatine (tCr) signal-to-noise (SNR), Water linewidth for SVS at both fields in two different locations. There were significant differences between field strength for SNR (*p* = 2.1× 10LJ¹²) and full width half maximum (FWHM) (*p* = 2.1× 10LJ¹^5^).

| Technique | tCr SNR | Water FWHM (ppm) | Field strength | Location |
| --- | --- | --- | --- | --- |
| SVS | $101.3 \pm 18$ | $0.043 \pm 0.0008$ | 3T | Lower limb |
| SVS | $99.7 \pm 24$ | $0.046 \pm 0.003$ | 3T | Upper limb |
| SVS | $148.0 \pm 10$ | $0.042 \pm 0.003$ | 7T | Lower limb |
| SVS | $132.7 \pm 17$ | $0.039 \pm 0.003$ | 7T | Upper limb |

#### 3.1.2. Reproducibility Comparison Between SVS and MRSI

Figure 6 presents the CV values calculated from two sessions for individual subjects (intra-subject) of SVS and MRSI techniques for metabolite concentration ratios to tCr across upper and lower limb (see Table S1, S2, and Figure S1 in supplementary for mean CV values). For MRSI, CV values ranged from 1.3% for tNAA/tCr at 3T (upper limb) to a maximum of 15.3% for Gln/tCr at 7T (lower limb), and, for SVS the smallest CV observed at 0.4% for tNAA/tCr 7T (upper limb) and the highest at 28.9% for Tau/tCr at 3T (upper limb). SVS demonstrated lower CV values for 5 out of 8 metabolites at 7T, indicating better reproducibility compared to MRSI. At 3T, MRSI showed lower CV values for 6 out of 8 metabolites compared to SVS. Field strength also influenced reproducibility, and MRSI shows better reproducibility at 3T compared to 7T for most metabolites. However, at 7T, SVS consistently provided better reproducibility, with lower CVs for most metabolites, including Glu/tCr and tNAA/tCr. Significant differences between the techniques were observed for certain metabolites. For example, at 7T in the upper limb, SVS showed significantly lower CVs for tNAA/tCr (*p* = 0.001) and Glx/tCr (*p* = 0.046) compared to MRSI.

**Figure 6:**
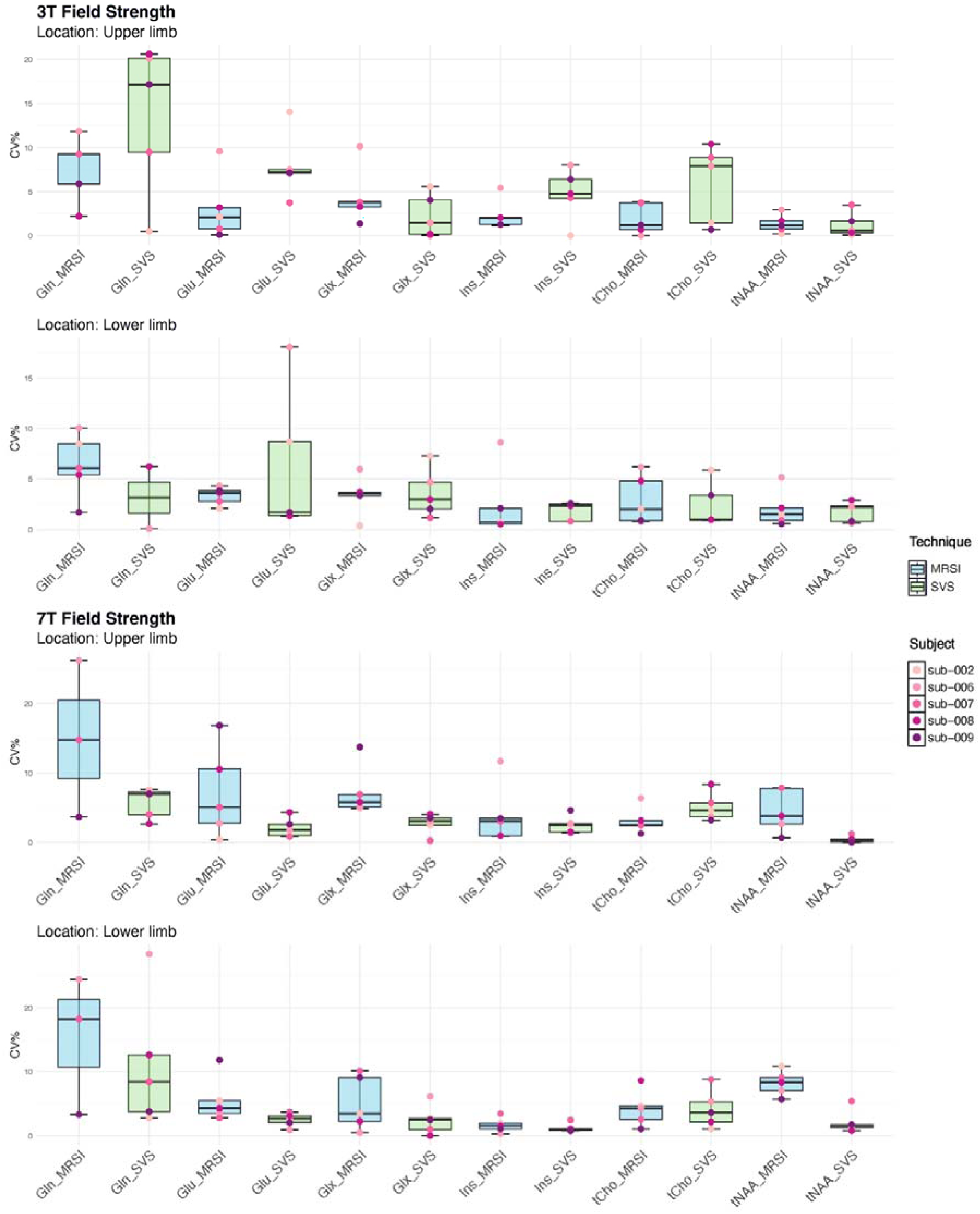
Intrasubject coefficient of variation (CV, %) between two sessions from five subjects for different metabolites/total creatine (tCr) in two brain regions at 3T (top) and 7T (bottom) using SVS-mask. MRSI is represented in blue and SVS in green. Most CVs are in the range of 0 and 10%, except for a few outliers. Some metabolites exhibit lower CVs means higher reproducibility than others, leading to a narrower value distribution and consequently smaller boxes in the box plot.

#### 3.1.3. Reproducibility Using GM and WM Segmentations Within SVS Mask

The results in Figure 7 demonstrated that GM generally exhibited higher mean CV values than WM for certain metabolites, such as Gln/tCr in lower limb GM at 7T (18.5%) compared to lower limb WM (15.6%). Furthermore, WM regions at 7T showed improved reproducibility, with lower mean CV values observed for most metabolites.

**Figure 7:**
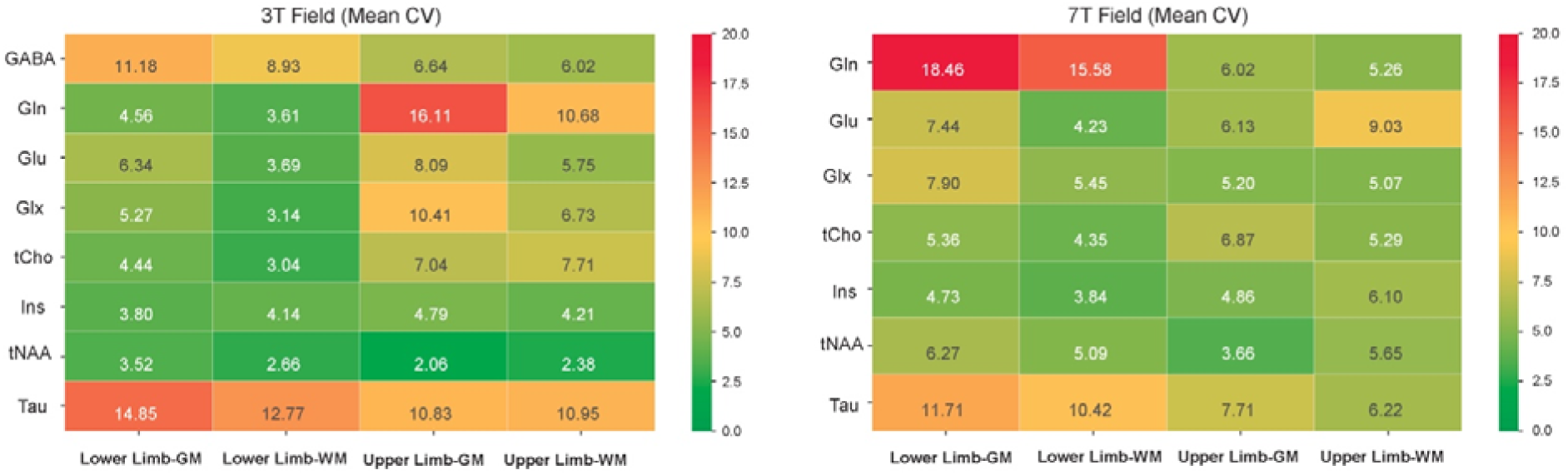
Heatmap of mean coefficient of variation (CV, %) for metabolite concentration ratios to total creatine (tCr) using grey matter (GM) and white matter (WM) segmentations within the SVS mask across 3T and 7T fields with 3D-CRT-FID-MRSI technique. The 3T field results (left) show generally higher CVs for GM compared to WM, particularly for metabolites such as glutamine (Gln). The 7T field results (right) reveal an improvement in reproducibility compared to 3T for WM regions, with lower CVs for most metabolites.

### 3.2. Evaluation of Region Averaging Approaches in MRSI

The correlation between “fit-first” and “average-first” approaches was assessed using a consistent WM segmentation within the SVS mask to reduce partial volume effects and selecting approximately 20-30 MRSI voxels to ensure sufficient SNR. Overall, the correlations between the two approaches were strong, ranging from “moderate” to “very high” across different B_0_ and metabolites^18,37^, with 7T generally showing higher correlations, such as Gln/tCr in the upper limb (r = 0.98, *p* < 0.001) and Glx/tCr (r = 0.95, *p* < 0.001). Although some metabolites, like tNAA/tCr at 7T, displayed lower correlations (r = 0.49, *p* = 0.14), differences between methods were largely non-significant (Figure 8).

**Figure 8:**
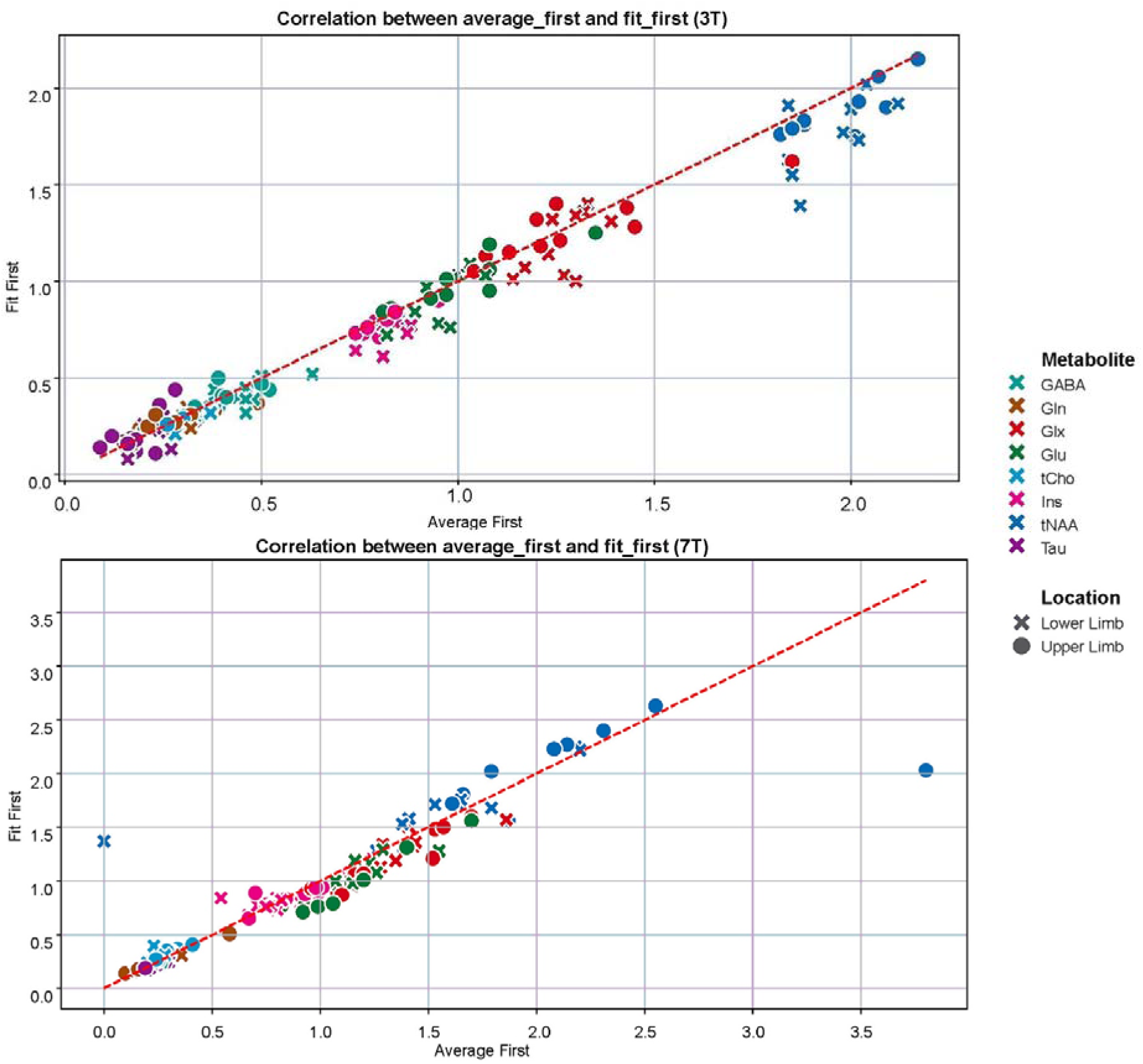
Scatter plot showing the correlation between “average-first” and “fit-first” metabolite concentration ratios at 3T (top) and 7T (bottom) for all measured metabolites. Each point represents the mean metabolite ratio to tCr, averaged across two sessions and five subjects for the upper limb and lower limb regions. The dashed red line indicates perfect agreement between the two techniques (“average-first” = “fit-first”). There are two outliers at 7T one for “fit-first” lower limb tNAA (too low) and one for “average-first” upper limb tNAA (too high).

In addition to the WM mask (Figure S2 in supplementary), atlas-based mask was also used to assess reproducibility of two region averaging approaches. In overall, both approaches demonstrated comparable reproducibility, with no significant differences in CV values for most metabolites (Figure S3 in supplementary). However, the “fit-first” approach generally yielded slightly lower CVs, indicating better reproducibility, particularly for metabolites such as tNAA/tCr and Glx/tCr at 7T in atlas-based regions. Notably, CVs were higher at 7T for most metabolites using both methods.

### 3.3. Voxel-Wise Correlation of MRSI Metabolite Measurements Across Sessions

The voxel-wise correlation analysis between MRSI measurements from two scanning sessions revealed varying degrees of reproducibility across different metabolites and B_0_. At 3T, strong positive correlations were observed for several metabolites, including tCho (r = 0.70) and tNAA (r = 0.47), indicating consistent metabolite quantification between sessions (Figure 9). However, metabolites like GABA and Gln showed lower correlation values, suggesting potential variability in these measurements. In contrast, at 7T, higher correlation coefficients were evident for most metabolites, such as tCho (r = 0.73), Glx (r = 0.63), and tNAA (r = 0.71) highlighting improved reproducibility at higher B_0_. Notably, Gln at 7T demonstrated moderate correlation (r = 0.51), while metabolite like Tau showed significant consistency with correlations exceeding 0.46.

**Figure 9:**
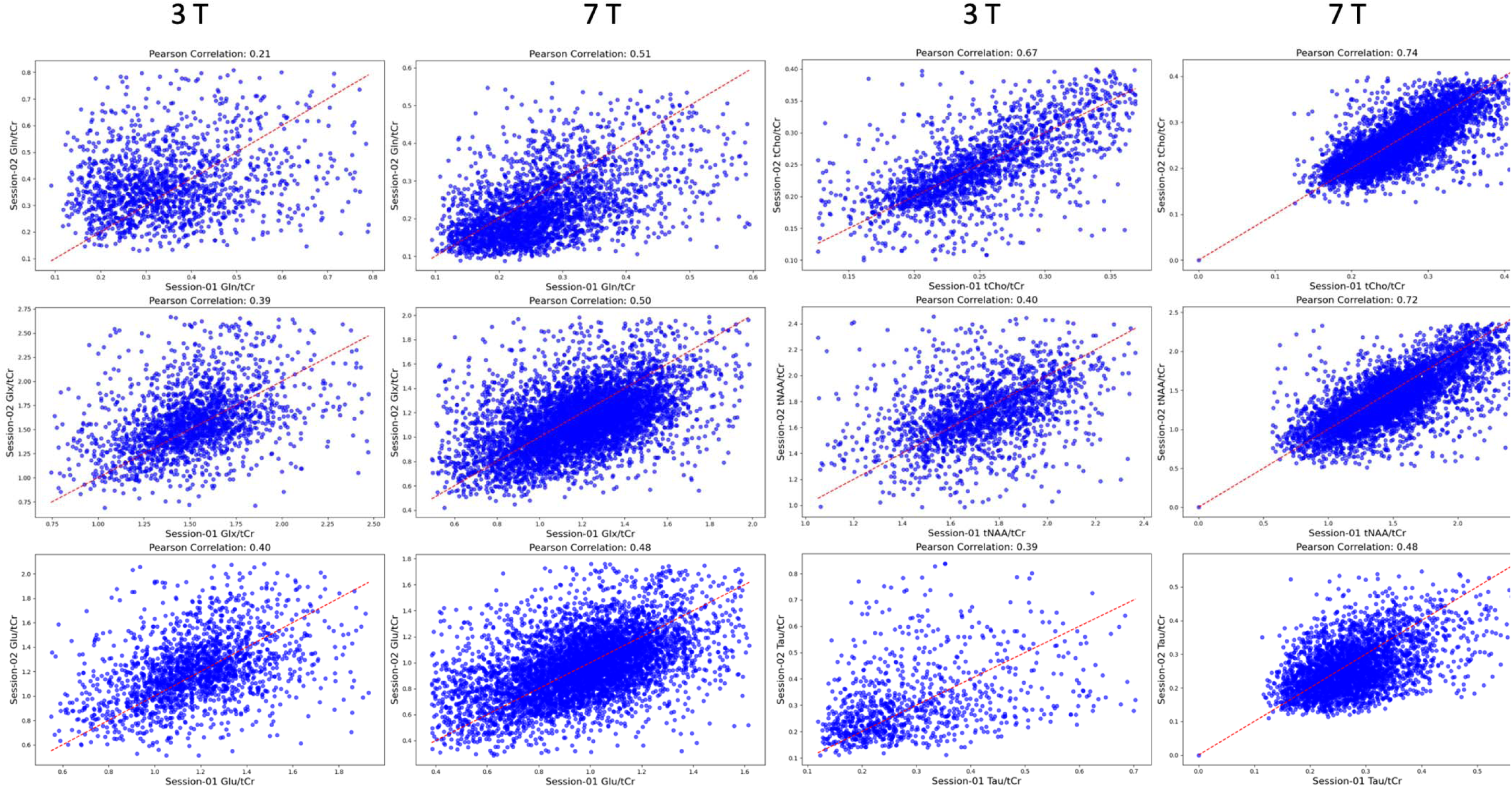
Scatter plots illustrating correlation of metabolite ratios across two sessions (session one vs. session two) for one subject as an example at both fields. Each plot represents a different metabolite ratio to total creatine (tCr) plotted for session one (x-axis) and session two (y-axis). The dashed red line represents the line of perfect agreement (y = x).

## 4. Discussion

SVS remains the most widely employed ^1^H-MRS technique for measuring brain metabolite concentrations, offering simplicity and availability on standard MR scanners. However, its limited spatial resolution and coverage constrain its application, particularly in studies requiring information from multiple brain regions. In contrast, the 3D-CRT-based FID-MRSI technique provides greater spatial coverage and resolution. These characteristics make MRSI preferred for capturing neuro-metabolic changes across the brain, particularly in clinical studies such as cancer and neurodegenerative diseases^11,61^. Before applying these methods to pathological conditions or treatment-induced neuro-metabolic changes, it is essential to evaluate their reproducibility in healthy participants.

### 4.1. Comparison with Literature

This study is the first to directly compare the reproducibility of SVS and MRSI in the same cohort of healthy participants at both 3T and 7T. Additionally, it introduces a unique analysis of two voxel spatial averaging approaches for MRSI “fit-first” and “average-first”. We also compare different masking strategies to highlight the unique capabilities of MRSI in isolating GM and WM regions. Good spectral quality was achieved for both techniques, and our concentration estimates were generally consistent with prior reports in the literature^11,18,19,37^. Our findings generally align with previous studies, although direct comparisons are challenging due to differences in acquisition protocols, field strengths, and spectroscopic sequences (Table S1 and S2 in supplementary)^18,37^. The high reproducibility (low CVs) observed in this study for both SVS and MRSI could be attributed to factors such as shorter scan times, smaller voxel sizes for SVS, number of subjects, and the inclusion of young, healthy participants^60,62,63^. For MRSI, our use of tailored ROIs, unique sequences, and different spatial averaging methods may have further contributed to the trend of lower CVs compared to prior studies^18,37,63-65^.

### 4.2. Reproducibility Across Techniques

The mean CV values for both SVS and MRSI indicated good-to-excellent reproducibility across most metabolites, with values ranging from 1.4% to 15.3% for MRSI and 0.4% to 28% for SVS. The reproducibility of low-concentration metabolites like Gln, GABA, and Tau varied significantly between the two techniques. For instance, MRSI demonstrated superior reproducibility for these metabolites in the upper limb, while SVS performed better in the lower limb region for certain ratios like Tau/tCr and GABA/tCr. While SVS is well-suited for studies focusing on single regions or when high spatial resolution is not essential, it faces challenges in evaluating small brain areas, such as the GM in the precentral gyrus and paracentral lobule, where poor B_0_ homogeneity and low SNR can affect data quality. MRSI, with its ability to capture metabolite profiles from a large section of the brain in one session, is advantageous in these contexts. Notably, some metabolites, such as NAA/tCr, displayed substantial variability between the two techniques of SVS and MRSI using SVS-based mask (mean CV range: 0.4% to 8.2%), which may be related to lipid contamination in specific regions or subjects. While SVS demonstrated lower CV values overall, the CV range observed for MRSI at both fields (1.4% to 15.3%) is still within an acceptable range for certain applications. For a technique like MRSI, which offers significantly broader spatial coverage compared to SVS, these values indicate promising reproducibility.

The reproducibility of MRSI at 7T was generally lower than at 3T, likely due to the longer scan duration (4:14 minutes at 3T vs. 12:11 minutes at 7T), which increases the risk of subject movement. Additionally, the real-time single-echo EPI navigators used at 3T allow for prospective motion and B0-field correction, helping to stabilize spectral quality and reduce intra-session variability. Since this correction method has not yet been successfully implemented at 7T, its absence may have further contributed to the higher CV values observed at 7T compared to 3T^37^. Moreover, the larger voxel sizes at 3T (6.4 times larger than at 7T) may have contributed to maintain SNR at a level comparable to 7T, compensating for the inherent SNR-penalty at 3T.

### 4.3. Evaluation of Region Averaging Approaches

A novel aspect of this study was the evaluation of two voxel averaging approaches in MRSI: “fit-first” and “average-first”. The rationale for the “average-first” approach lies in its potential to improve the signal for low-concentration metabolites like Tau, Gln, and GABA before the fit is performed. By aligning and averaging signals from multiple voxels, random noise cancels out while coherent metabolite signals constructively add. This results in a cleaner spectrum with sharper peaks, facilitating more accurate metabolite quantification. This method, however, relies on high data quality, as poor-quality voxels within the mask can disproportionately affect the results. Unlike the “fit-first” approach, where individual voxels can be examined and potentially excluded after fitting, the “average-first” approach does not allow for the identification of specific voxel outliers before averaging, as the process produces a single spectrum for the entire ROI. Future work should focus on improving outlier detection methods to enhance the “average-first” approach by identifying and removing poor-quality voxels before averaging. Our results showed a strong correlation between the two approaches, with significant differences in reproducibility for certain metabolite concentration ratios in specific regions (Figure S2 supplementary). The “fit-first” approach offers a practical advantage in this context, allowing researchers to identify and exclude (a small amount of) outlier voxels, thereby enhancing the reliability of ROI-based analyses. In contrast, the “average-first” approach can produce inaccurate results if even a small number of voxels are of poor quality, as these cannot be identified and excluded post-averaging.

### 4.4. Features of MRSI

MRSI offers unique capabilities unattainable with SVS, such as evaluating metabolite concentration ratios exclusively within GM or WM. CV values indicated higher variability in GM compared to WM, particularly at 7T for metabolites like Gln (in lower limb 18.46% in GM vs. 15.58% in WM). This finding is somewhat unexpected, as Gln concentration is generally higher in GM than in WM, which should theoretically result in a higher SNR and more reliable spectral fitting^66^. Additionally, WM regions at 7T displayed lower CVs for most metabolites, indicating better reproducibility, higher homogeneity of WM tissue and reduced susceptibility to partial volume effects^67^. These findings highlight MRSI’s advantage in spatial resolution and its ability to extract region-specific metabolic information, which is crucial for understanding the neurochemical basis of diseases. Unlike SVS, where the target region must be selected before acquisition, MRSI allows for retrospective analysis, enabling researchers to define ROIs post hoc. This flexibility is particularly valuable in studies where the most relevant brain regions are not known beforehand or when new insights emerge during data analysis.

### 4.5. Limitations

Firstly, the sample size was smaller compared to other reproducibility studies, primarily due to the extensive scanning protocol, which required approximately 5 hours per participant across multiple sessions. These sessions included scans at two field strengths, using two different sequences and voxel locations, which significantly limited the number of participants that could be included. Secondly, at 7T we attempted to mitigate motion by selecting younger participants and using padding to restrict movement, because real-time motion correction has not been implemented at 7T yet and we used real-time correction only at 3T, which has been demonstrated to enhance data quality in high-resolution 3D-CRT-based FID-MRSI at 3T^37^. Its application at 7T, especially given the longer scan time of 12 minutes, could have further improved data quality at 3T. Thirdly, the scanning protocols were tailored to optimize the performance of each technique, providing good spectral quality within a reasonable measurement time. However, matching acquisition parameters (e.g., TR and TE) between SVS and MRSI for direct comparison was not feasible due to structural and timing differences in the sequences, such as differing minimum TEs. Fourthly, the choice of ROIs was restricted to the upper and lower limb regions of the motor area, based on prior research and the specific interests of this laboratory. This limits the generalizability of findings to other brain regions. Finally, the last limitation was the absence of a water reference scan for the MRSI sequence. Without this, the estimate metabolite concentrations were not possible, and referencing to an internal standard (tCr) was used instead without T_1_ and T_2_ corrections.

## 5. Conclusion

Our results suggest that the ratio of metabolite concentrations to tCr can be created with both sLASER and 3D-CRT-FID-MRSI with a good to excellent reproducibility at both 3T and 7T for most brain regions assessed. While SVS offers superior reproducibility in certain contexts, MRSI’s spatial coverage and ability to isolate GM and WM make it a valuable approach for comprehensive neurochemical mapping with good reproducibility. The comparison of region averaging approaches for MRSI highlights the importance of methodological choices in achieving reliable results, particularly for low-concentration metabolites. Scan time seems to be an important factor for having reproducible results, and one could test to reduce scan time at 7T if lower spatial resolution is not an issue. Future studies should continue to explore these techniques in pathological conditions to better understand their clinical relevance.

## Abbreviations used

Cr: creatine
CRLB: Cramér-Rao lower bound
CRT: concentric ring trajectory
CSDE: chemical shift displacement error
CSF: cerebrospinal fluid
CV: coefficient of variation
FID: free induction decay
FWHM: full width at half maximum
GABA: γ-aminobutyric acid
Gln: glutamine
Glu: glutamate
Glx: Glu + Gln
GM: grey matter
GOIA: gradient offset independent adiabatic
GPC: glycerophosphocholine
HC: healthy control
myo-Ins: myo-inositol
MND: motor neuron disease
MRSI: magnetic resonance spectroscopic imaging
NAA: *N*-acetylaspartate
NAAG: *N*-acetylaspartylglutamate
OVS: outer volume suppression
PCho: ghosphorylcholine
PCr: phosphocreatine
ppm: parts per million
ROI: region of interest
SAR: specific absorption rate
sLASER: semi-localization by adiabatic selective refocusing
SNR: signal-to-noise ratio
SVS: single voxel spectroscopy
Tau: taurine
tCho: total choline (GPC + PCho)
tCr: total creatine (Cr + PCr)
TE: echo time
TR: repetition time
tNAA: total NAA (NAA + NAAG)
UHF: ultra-high field
VAPOR: variable pulse power and optimised relation delays
VOI: volume of interest
WET: water suppression enhanced through T1 effects
WM: white matter

## 6. Acknowledgements

We acknowledge funding by the Australian Research Council Training Centre for Innovation in Biomedical Imaging Technology (IC170100035). TBS is supported by a Motor Neurone Disease Research Australia (MNDRA) Postdoctoral Research Fellowship (PDF2112) and NHMRC Ideas grant APP2029871. FN is supported by Austrian Science Fund (project number 10.55776/KLI1106). WB is supported by the National Institutes of Health grants: R01EB031787. The author used ChatGPT for basic linguistic checking for some parts of this paper. The authors acknowledge the facilities of the National Imaging Facility at the Centre for Advanced Imaging. We thank our research radiographers, Nicole Atcheson, Aiman Al-Najjar and Sarah Daniel for assisting in the data collection. We thank Dr. Georg Oeltzschner for helping to set up pipeline for postprocessing of MRS data and MRS hub website. We are grateful to the volunteers who participated in this study. Open access publishing facilitated by The University of Queensland, as part of the Wiley - The University of Queensland agreement via the Council of Australian University Librarians. The MRS package was developed by Gülin Öz and Dinesh Deelchand for the semi-LASER sequence and the basis set used for spectral fitting described in ^29,68,69^; and provided by the University of Minnesota under a C2P agreement. 3D-CRT-FID-MRSI sequence and the basis sets for 3T and 7T were provided by the high-field magnetic resonance centre (HFMRC) at the Medical University of Vienna.

