## Supplementary Material for "From a Voxel to Maps: A Comparative Study of sLASER SVS and 3D-CRT-FID-MRSI at 3 T and 7 T"

### 7. Supplementary Materials

Table S1: Mean (std) values of metabolite concentration to tCr ratio and mean CV in lower limb region for both techniques at 3T and 7T

| Metabolite | Field | Technique | Mean (SD) | Mean CV ± std |
|---|---|---|---|---|
| **GABA/tCr** | 3T | MRSI | 0.45 (0.07) | 10.2 ± 6.5 |
| | 7T | MRSI | 0.31 (0.10) | NA (excluded) |
| | 3T | SVS | Nan (excluded) | NA (excluded) |
| | 7T | SVS | Nan (excluded) | 24.6 ± 21.7 |
| **Gln/tCr** | 3T | MRSI | 0.35 (0.04) | 6.4 ± 3.2 |
| | 7T | MRSI | 0.24 (0.07) | 15.3 ± 10.8 |
| | 3T | SVS | 0.13 (0.04) | 3.2 ± 9.4 |
| | 7T | SVS | 0.26 (0.05) | 11.2 ± 12.8 |
| **Glu/tCr** | 3T | MRSI | 1.07 (0.09) | 3.3 ± 0.9 |
| | 7T | MRSI | 1.08 (0.19) | 5.6 ± 3.6 |
| | 3T | SVS | 0.92 (0.09) | 6.2 ± 3.3 |
| | 7T | SVS | 0.91 (0.04) | 2.5 ± 1.8 |
| **Glx/tCr** | 3T | MRSI | 1.42 (0.11) | 3.4 ± 2.0 |
| | 7T | MRSI | 1.21 (0.18) | 5.1 ± 4.3 |
| | 3T | SVS | 1.10 (0.13) | 3.6 ± 2.0 |
| | 7T | SVS | 1.18 (0.08) | 2.5 ± 3.7 |
| **Ins/tCr** | 3T | MRSI | 0.84 (0.04) | 2.5 ± 3.5 |
| | 7T | MRSI | 0.85 (0.07) | 1.7 ± 1.2 |
| | 3T | SVS | 0.86 (0.03) | 1.8 ± 4.2 |
| | 7T | SVS | 0.80 (0.03) | 1.3 ± 2.7 |
| **Tau/tCr** | 3T | MRSI | 0.26 (0.06) | 14.7 ± 11.2 |
| | 7T | MRSI | 0.23 (0.03) | 6.5 ± 1.4 |
| | 3T | SVS | 0.19 (0.05) | 19.1 ± 18.0 |
| | 7T | SVS | 0.20 (0.04) | 9.7 ± 7.6 |
| **tCho/tCr** | 3T | MRSI | 0.32 (0.03) | 2.9 ± 2.4 |
| | 7T | MRSI | 0.32 (0.04) | 4.3 ± 2.8 |
| | 3T | SVS | 0.27 (0.03) | 2.4 ± 1.6 |
| | 7T | SVS | 0.21 (0.03) | 4.2 ± 1.5 |
| **tNAA/tCr** | 3T | MRSI | 1.93 (0.09) | 2.1 ± 1.8 |
| | 7T | MRSI | 1.74 (0.27) | 8.2 ± 2.0 |
| | 3T | SVS | 1.47 (0.05) | 1.8 ± 1.3 |
| | 7T | SVS | 1.65 (0.08) | 2.2 ± 1.8 |

Table S2: Mean (std) values of metabolite concentration to tCr ratio and mean CV in upper limb region for both techniques at 3T and 7T

| Metabolite | Field | Technique | Mean (SD) | Mean CV ± std |
|---|---|---|---|---|
| GABA/tCr | 3T | MRSI | 0.43 (0.05) | 6.5 ± 3.8 |
| | 7T | MRSI | 0.31 (0.06) | NA (excluded) |
| | 3T | SVS | 0.13 (0.03) | NA (excluded) |
| | 7T | SVS | 0.12 (0.04) | 5.8 ± 8.2 |
| Gln/tCr | 3T | MRSI | 0.33 (0.04) | 7.7 ± 3.7 |
| | 7T | MRSI | 0.25 (0.13) | 14.9 ± 11.2 |
| | 3T | SVS | 0.28 (0.05) | 13.6 ± 9.6 |
| | 7T | SVS | 0.24 (0.04) | 5.7 ± 6.7 |
| Glu/tCr | 3T | MRSI | 1.05 (0.12) | 3.2 ± 3.8 |
| | 7T | MRSI | 1.10 (0.20) | 7.1 ± 6.6 |
| | 3T | SVS | 1.02 (0.09) | 7.9 ± 3.4 |
| | 7T | SVS | 0.90 (0.03) | 2.1 ± 2.9 |
| Glx/tCr | 3T | MRSI | 1.37 (0.15) | 4.5 ± 3.3 |
| | 7T | MRSI | 1.23 (0.24) | 7.3 ± 3.7 |
| | 3T | SVS | 1.30 (0.07) | 2.3 ± 1.6 |
| | 7T | SVS | 1.14 (0.07) | 2.7 ± 3.7 |
| Ins/tCr | 3T | MRSI | 0.82 (0.03) | 2.4 ± 1.8 |
| | 7T | MRSI | 0.87 (0.13) | 4.0 ± 4.4 |
| | 3T | SVS | 0.83 (0.04) | 4.7 ± 2.9 |
| | 7T | SVS | 0.78 (0.02) | 2.6 ± 3.5 |
| Tau/tCr | 3T | MRSI | 0.24 (0.07) | 9.9 ± 8.2 |
| | 7T | MRSI | 0.23 (0.03) | 4.6 ± 0.9 |
| | 3T | SVS | 0.15 (0.03) | 28.9 ± 27.3 |
| | 7T | SVS | 0.21 (0.03) | 9.8 ± 9.5 |
| tCho/tCr | 3T | MRSI | 0.31 (0.02) | 1.9 ± 1.8 |
| | 7T | MRSI | 0.33 (0.05) | 3.1 ± 1.9 |
| | 3T | SVS | 0.26 (0.03) | 5.8 ± 2.6 |
| | 7T | SVS | 0.22 (0.03) | 5.1 ± 2.2 |
| tNAA/tCr | 3T | MRSI | 1.96 (0.09) | 1.4 ± 1.0 |
| | 7T | MRSI | 1.99 (0.32) | 4.6 ± 3.2 |
| | 3T | SVS | 1.70 (0.09) | 1.2 ± 1.4 |
| | 7T | SVS | 1.83 (0.08) | 0.4 ± 1.0 |

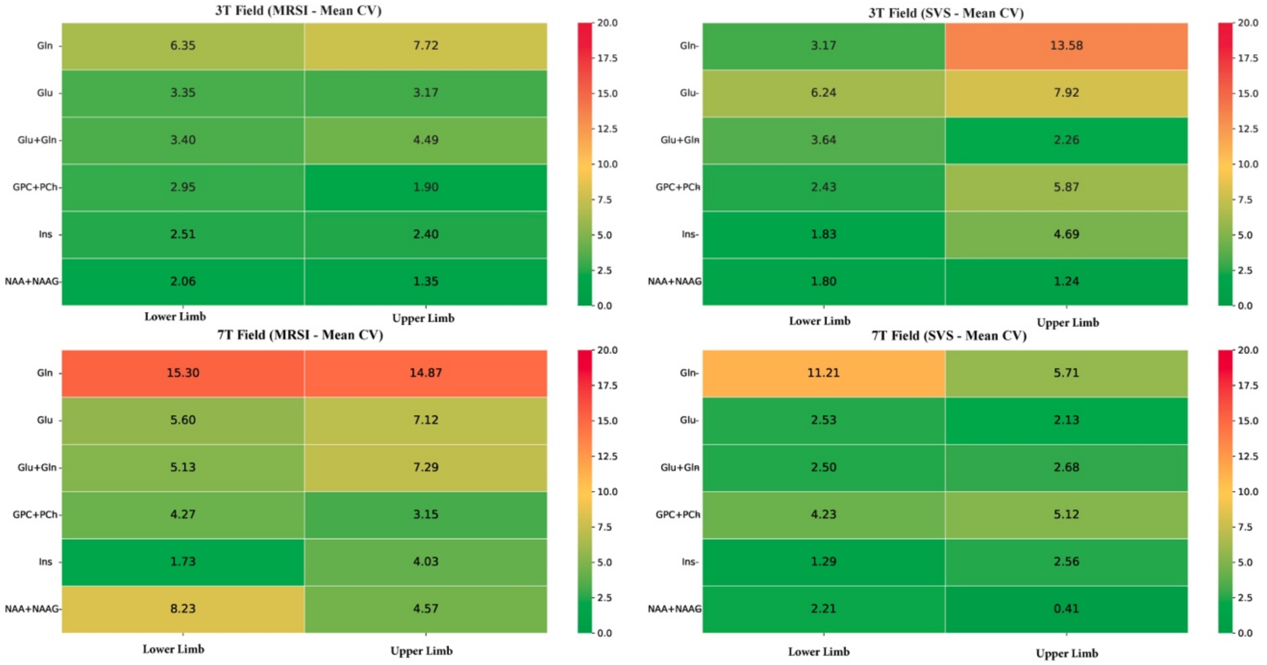


Figure S1: The heatmap illustrates the mean CV distributions across metabolites/tCr, using the SVS-based masking for both MRSI and SVS techniques in two region and two field. This comparison highlights the reproducibility of the two techniques with higher CV values indicating poorer reproducibility.

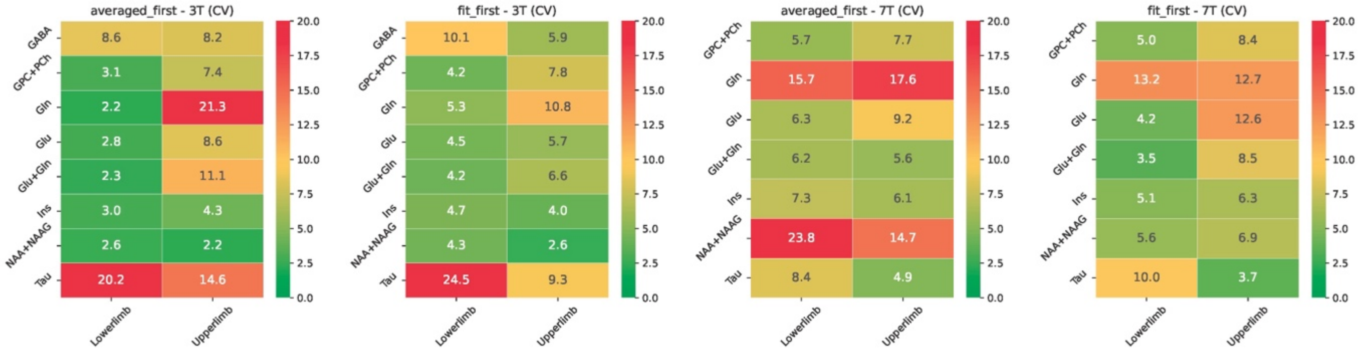


Figure S2: The heatmap illustrates the CV distributions across metabolites/tCr, focusing on a small region of WM within the SVS mask using 3D-CRT-FID-MRSI technique at both fields. This comparison highlights the reproducibility of the two spatial averaging approaches ("fit-first" and "average-first"), with higher CV values (red) indicating poorer reproducibility.

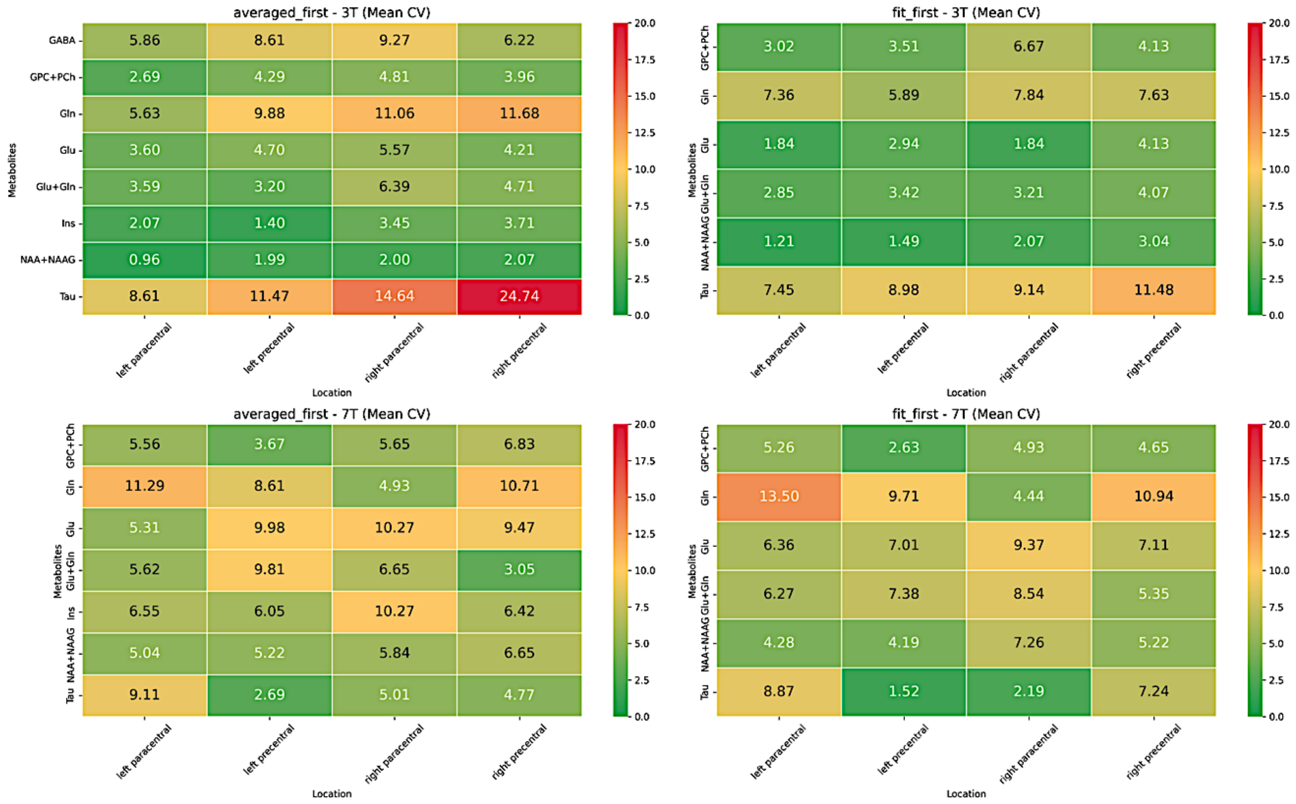


Figure S3: The heatmap illustrates the CV distributions across metabolites/tCr, focusing on Atlas_based masking using 3D-CRT-FID-MRSI technique at both fields. This comparison highlights the reproducibility of the two spatial averaging approaches ("fit-first" and "average-first"), with higher CV values indicating poorer reproducibility.

Table S3. THE MRSinMRS CHECKLIST

| | | | | |
|---|---|---|---|---|
| **1. Hardware** | | | | |
| **a. Field strength (T)** | 3 | | 7 | |
| **b. Manufacturer** | Siemens | | Siemens | |
| **c. Model** | ve syngo MR E11 | | vd syngo MR E12 | |
| **d. RF coils** | 62 ch $^1$H head coil | | 32 ch $^1$H head coil | |
| **2. Acquisition** | | | | |
| **a. Pulse sequence** | CRT-FID-MRSI | sLASER | CRT-FID-MRSI | sLASER |
| **b. VOI locations** | Cerebrum | Precentral Gyrus and Paracentral Lobule | Cerebrum | Precentral Gyrus and Paracentral Lobule |
| **c. Voxel size (mm$^3$)** | 6.3×6.3×6.2 | 25×25×25 | 3.4×3.4×3.5 | 25×25×25 |
| **d. TR/TE (ms)** | 950/0.8 | 2000/28 | 460/1.3 | 8000/26 |
| **e. Averages** | 1 | 64 | 1 | 16 |

| | | | | |
|---|---|---|---|---|
| **f. spectral width in Hz, number of spectral points; If MRSI: 2D or 3D, FOV in all directions, matrix size, acceleration factors, sampling method** | BW 1325, MRSI: 3D, 200×200×130 mm3, 32×32×21, spatial-spectral encoding | BW 4000, 2048 spectral points | BW 2778, MRSI: 3D, 220×220×110 mm3 , 64×64×31, spatial-spectral encoding | BW 6000, 2048 spectral points |
| **g. Water suppression method** | WET | VAPOR | WET | VAPOR |
| **h. Shimming method** | Standard shim + manual adjustment, water peak < 50 Hz | fastestmap | Standard shim + manual adjustment, water peak < 50 Hz | fastestmap |
| **3. Data analysis methods and outputs** | | | | |
| **a. Analysis software** | LCModel 6.3-1 | Osprey | LCModel 6.3-1 | Osprey |
| **b. Processing steps deviating from quoted reference or product** | Internal water reference | Default Osprey | Internal water reference | Default Osprey |
| **c. Output measure** | Ratio/tCr | | | |
| **d. Quantification references and assumptions, fitting model assumptions** | Simulated in NMRScope-B, macromolecular background | Basis set includes 19 simulated metabolites + measured macromolecule. LCModel basline knot spacing 5.00 ppm | Simulated in NMRScope-B, macromolecular background | Basis set includes 19 simulated metabolites + measured macromolecule. LCModel basline knot spacing 5.00 ppm |
| **4. Data quality** | | | | |
| **a. Reported variables (SNR, linewidth (with reference peaks))** | MRSI: SNR for NAA was estimated voxel-wise using the pseudo-replica method, which incorporates receiver noise prescans acquired at the start of the MRSI sequence. The FWHM of the spectral peaks for tCr at 3.02 ppm was calculated from the LCModel fits. sLASER: SNR dividing the tCr by the standard deviation of noise within the range of -2 to 0 ppm. Linewidth FWHM of a Lorentzian peak model for the water peak between 4.4 and 5.0 ppm | | | |
| **b. Data exclusion criteria** | CRLB > 50% | | | |

| | |
|---|---|
| **c. Quality measures of postprocessing model fitting (eg CRLB, goodness of fit, SD of residual)** | CRLB, and SD of residual |
| **d. Sample spectrum** | Figure 2 |
